# Impact of the COVID-19 pandemic on the malaria burden in northern Ghana: Analysis of routine surveillance data

**DOI:** 10.1101/2021.11.29.21266976

**Authors:** Anna-Katharina Heuschen, Alhassan Abdul-Mumin, Martin Nyaaba Adokiya, Guangyu Lu, Albrecht Jahn, Oliver Razum, Volker Winkler, Olaf Müller

## Abstract

**Introduction:** The COVID-19 pandemic and its collateral damage severely impact health systems globally and risk to worsen the malaria situation in endemic countries. Malaria is a leading cause of morbidity and mortality in Ghana. This study aims to analyze routine surveillance data to assess possible effects on the malaria burden in the first year of the COVID-19 pandemic in the Northern Region of Ghana.

**Methods:** Monthly routine data from the District Health Information Management System II (DHIMS2) of the Northern Region of Ghana were analyzed. Overall outpatient department visits and malaria incidence rates from the years 2015 to 2019 were compared to the corresponding data of the year 2020.

**Results:** Compared to the corresponding periods of the years 2015 to 2019, overall visits and malaria incidence in pediatric and adult outpatient departments in northern Ghana decreased in March and April 2020, when major movement and social restrictions were implemented in response to the pandemic. Incidence slightly rebounded afterwards in 2020 but stayed below the average of the previous years. Data from inpatient departments showed a similar but more pronounced trend when compared to outpatient departments. In pregnant women, however, malaria incidence in outpatient departments increased after the first COVID-19 wave.

**Discussion:** The findings from this study show that the COVID-19 pandemic affects the malaria burden in health facilities of Ghana, with declines in in- and outpatient rates. Pregnant women may experience reduced access to intermittent preventive malaria treatment and insecticide treated nets, resulting in subsequent higher malaria morbidity. Further data from other African countries, particularly on community-based studies, are needed to fully determine the impact of the pandemic on the malaria situation.

## Introduction

Malaria remains one of the leading causes of morbidity and mortality in sub-Saharan Africa (SSA). It is responsible for nearly one quarter of all under five childhood deaths in this region (1, 2).

The global spread of the coronavirus disease 2019 (COVID-19) was declared a Public Health Emergency of International Concern, which is the highest level of alarm, at the end of January 2020 (3). Many African governments responded rapidly to this threat by implementing control measures even before first cases were detected in their countries, comprising border closures, movement restrictions, social distancing and school closures (4). By November 2021, there were nearly 6.2 million COVID-19 cases reported from the WHO African Region, with about 152,000 deaths, mostly from the southern and northern rims of the continent (5). In the global context, SSA accounts for only about 2.5% and 3% of the overall reported COVID-19 morbidity and mortality, respectively, while it is home to 17% of the global population (6-8). This may be explained by factors such as a younger population, hotter climate, interferences with other infectious diseases, and especially lack of diagnostics and underreporting (9, 10). Ghana is among the countries with the highest reported COVID-19 cases (130,920) and deaths (1,209) in western and central SSA, as of November 2021 (8). COVID-19 vaccinations started in February 2021 but coverage in Ghana is still low with only 2.7% of the population fully vaccinated by November 2021 (11).

The socio-economic disruptions associated with the disease and the preventive measures present huge challenges for health systems and whole societies, especially in low- and middle income countries (12). In the highly malaria-endemic African countries, the progress made in malaria control during the last two decades is feared to be reversed by the side effects of the COVID-19 pandemic (13, 14).

This study aims to compare the malaria burden in the Northern Region of Ghana in the first year of the pandemic to previous years to assess whether a reversal indeed occurred.

## Methods

### Study area

Ghana, with its population of about 31 million, lies in western SSA and has a relatively well functioning health care system (15, 16). Ghana is divided into 16 administrative regions. The Northern Region, with its capital city Tamale, had a population of 1.9 million in 2020. The socio-economic situation of the Northern Region is below the national average of the country and the region has the highest rate of mortality under the age of five years (17). The rainy season in northern Ghana, which is usually associated with an increase in the malaria incidence, lasts from May to October (18).

Malaria is highly endemic in Ghana; the country accounts for 2% of the global malaria morbidity and 3% of the malaria mortality (19, 20). In 2020, malaria was the cause of 34% of all outpatient attendances (21). Treatment expenditures for common diseases like malaria are covered by a health insurance (22).

The first two confirmed COVID-19 cases in Ghana were seen on March 12, 2020; two days later, all public gatherings were banned. Travel restrictions and border closures were implemented on March 22, 2020 and the country’s major cities were placed under partial lockdown soon after. Schools were partially reopened on June 21, 2020 and borders were reopened to international airlines on September 21, 2020 (23). In Ghana, effects of the COVID-19 pandemic on malaria control interventions concerned the country’s stock of artemisinin-based combination therapies (ACT), the functioning of its insecticide-treated mosquito net (ITN) routine distribution, and the overall access to primary health care services and facilities (24).

### Study design and data

This retrospective observational study uses monthly malaria morbidity data on the overall number of outpatients (interpreted as less severe cases) and inpatients (more severe cases). Additionally, all outpatient visits (including non-malaria related visits) are analyzed. Cases were extracted from the district health information management system II (DHIMS2) on demographic and health parameters of northern Ghana from January 1, 2015, to December 31, 2020. This system was implemented in 2007 with an update in 2012 and has improved the data quality and completeness since (25).

Malaria diagnosis was based either on the results of rapid diagnostic tests or microscopy. Mid-year population estimates of the Northern Region of Ghana were also provided through the DHIMS2.

### Analysis

The data have been processed with Microsoft Excel Version 16.52 and analyzed with Stata IC Version 16 (Statacorp, College Station, TX, USA). We have calculated and plotted monthly incidence rates of all outpatient visits and confirmed malaria cases for the year 2020 and as a comparison for the years 2015 to 2019 separately and combined using population figures of the Northern Region of Ghana. Additionally, we calculated incidence rate ratios with 95% confidence intervals (95% CI) comparing quarterly incidence rates of 2020 versus the combined rates of 2015 to 2019. The data allowed analyzing children under five years and pregnant women separately using the fraction of the under-five population (14% of the population) and the fraction of women between 15 and 45 years (23% of the population) as estimates of the respective population denominators (26).

## Results

Table 1 presents a brief description of the dataset. Altogether 5.8 million outpatient department visits were reported between 2015 and 2020; 39% of those included a malaria diagnosis. Of all confirmed malaria cases, 20% were children under the age of five years and 2% were pregnant women. 295,465 patients were hospitalized with diagnosed malaria, 56% of those were children under the age of five years. The mean population of the years from 2015 to 2020 was 1,842,701 with 14% of children under the age of five years and 23% of women between the age of 15 and 45 considered as of possible childbearing age.

**Table 1:** Description of the dataset

|  | Number | Percentage (%) |
| --- | --- | --- |
| outpatient department visits |  |  |
| All | 5,804,910 | 100 |
| Malaria confirmed | 2,278,296 | 39 |
| Malaria confirmed among children <5 years | 454,779 | 20 |
| Malaria confirmed among pregnant women | 46,693 | 2 |
| hospital-admitted patients |  |  |
| Malaria confirmed | 295,465 | 100 |
| Malaria confirmed among children <5 years | 165,313 | 56 |
| mean mid-year population |  |  |
| Total population | 1,842,701 | 100 |
| Children <5 years* | 257,978 | 14 |
| Women aged 15 to 45* | 423,821 | 23 |

Figure 1 presents the incidence rates of the different outcomes reported in the Northern Region of Ghana for the years between 2015 and 2020 separately as well as a combined rate for the period 2015 to 2019. All visits of the outpatient department (OPD) (see Figure 1a), including also non-malaria patients, have experienced a major decline in March/April 2020, the months where COVID-19 control measures were implemented in the country, and stayed low during the following months. After a further decrease in September 2020, the numbers increased again in October 2020 to the levels observed in previous years. This trend is similar but not as pronounced in the general malaria OPD visits (Figure 1b). In children under the age of five years, the decline in accessing OPD malaria health care is stronger, especially from June to September 2020 (Figure 1 c). In pregnant women, however, a different trend with an earlier increase, starting in June and exceeding previous year’s levels, can be observed (Figure 1d). The 2020 numbers of the hospitally admitted malaria patients stayed below the previous standards from March to October 2020 (Figure 1e); and in accordance with the OPD figures, this trend is more pronounced in the children under five years population (Figure 1f).

**Figure 1:**
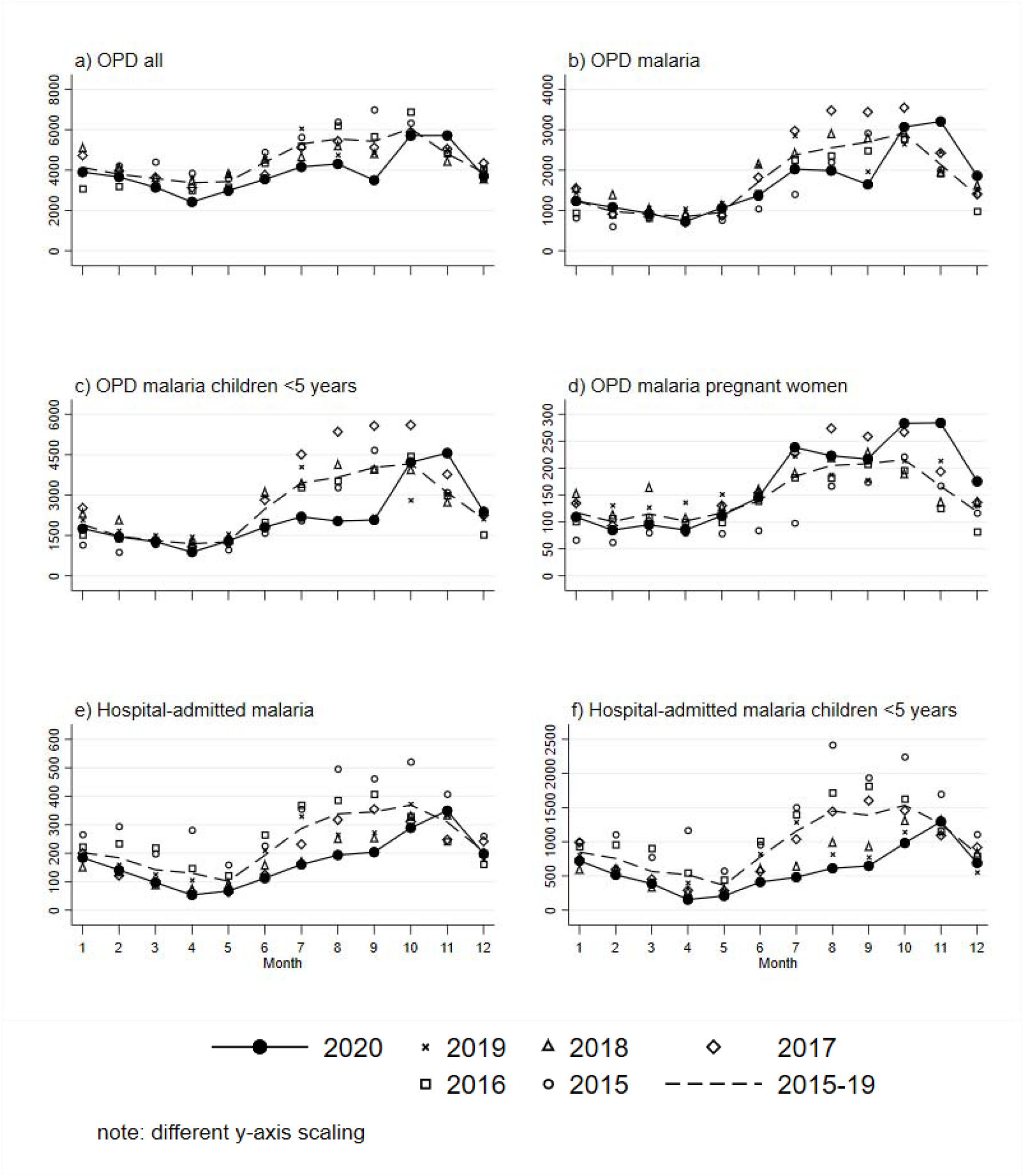
Reported monthly incidence rates per 100,000 of the Northern Region, Ghana for the years 2015 to 2020

Incidence rate ratios (IRR) depicting quarterly measures comparing the rates of 2020 to the combined rate of the years 2015 to 2019 are presented in table 2. General OPD visits were reduced in the 2^nd^ and 3^rd^ quarters of 2020 compared to the previous years (IRR 3^rd^ quarter 0.736) with a return to previous standards at the end of the year. The same applies to the overall malaria cases (IRR 0.742 in the 3^rd^ quarter) but with increases in the 4^th^ quarter (IRR 1.265). Ambulatory malaria cases in children under five experienced stronger reductions compared to previous years with an IRR 0.566 in the 3^rd^ quarter of 2020. These evolutions are not mirrored by the population of pregnant women with malaria infections, where no major reductions were observed during the first quarters of 2020 compared to previous years but with an earlier increase (IRR 1.481 in the 4^th^ quarter). The situation is slightly different in malaria infected patients admitted to the hospital. The reductions in the 2^nd^ and 3^rd^ quarters of 2020 are more pronounced (IRR 0.548 for all ages in the 2^nd^ quarter) and the numbers do not fully recover at the end of the year. Again, as for the outpatient population, this trend is more pronounced in children under five years of age (IRR 0.465 in the 2^nd^ quarter).

**Table 2:** Quarterly incidence rate ratios (IRR) with 95% confidence intervals (95% CI) comparing the incidence rates of 2020 with the combined incidence rates of the years 2015 to 2019

| Outcome | IRR (95% CI)<br>1<sup>st</sup> Quarter | IRR (95% CI)<br>2<sup>nd</sup> Quarter | IRR (95% CI)<br>3<sup>rd</sup> Quarter | IRR (95% CI)<br>4<sup>th</sup> Quarter |
| --- | --- | --- | --- | --- |
| outpatient department visits |  |  |  |  |
| All | 0.930<br>(0.925-0.934) | 0.800<br>(0.796-0.804) | 0.736<br>(0.732-0.739) | 1.026<br>(1.022-1.030) |
| Malaria | 1.035<br>(1.026-1.044) | 0.899<br>(0.892-0.907) | 0.742<br>(0.737-0.746) | 1.265<br>(1.258-1.272) |
| Malaria children <5 years | 0.956<br>(0.937-0.974) | 0.806<br>(0.790-0.823) | 0.566<br>(0.557-0.575) | 1.190<br>(1.176-1.206) |
| Malaria pregnant women | 0.865<br>(0.815-0.918) | 0.957<br>(0.905-1.011) | 1.136<br>(1.091-1.182) | 1.481<br>(1.424-1.540) |
| hospital-admitted patients |  |  |  |  |
| Malaria | 0.799<br>(0.780-0.817) | 0.548<br>(0.531-0.565) | 0.574<br>(0.563-0.586) | 0.946<br>(0.930-0.962) |
| Malaria children <5 years | 0.749<br>(0.726-0.773) | 0.465<br>(0.445-0.486) | 0.435<br>(0.422-0.448) | 0.820<br>(0.800-0.839) |

## Discussion

Since the beginning of the COVID-19 pandemic, several modelling studies have predicted negative collateral effects on the malaria burden in SSA, considering especially disrupted ITN campaigns and a limited access to antimalarial drugs. The study team of Weiss et al. created nine scenarios for different reductions of ITN coverage and access to antimalarial medication as well as regarding effects on malaria morbidity and mortality. As no ITN mass campaigns were scheduled for 2020 in Ghana, the worst-case scenario would have been a decline in access to antimalarials by 75% resulting in an increase of malaria morbidity and mortality by 12.6% and 54.6%, respectively (13). Overall, the predicted public health relevant effects of the COVID-19 pandemic on malaria include shared clinical disease manifestations leading to diagnostical challenges, disruptions of the availability of curative and preventive malaria commodities, significant effects on malaria programs, and in particular reduced access to malaria health services and health facilities in general (27).

In this study, we observed a slight but significant decline in malaria incidence during the 2^nd^ and 3^rd^ quarter of 2020 (April to September), and only a rebound to the average levels of previous years at the end of 2020. This pattern was visible in both, outpatient and inpatient settings, but more pronounced in the hospitalized population. The same applies to children and adults, where the reductions were also observed in both groups, but were more marked in children under five years of age. The marked decline in March/April 2020 can be explained by the extensive restrictions of movement and gathering and early stay-at-home advices for COVID-19-like symptoms unless these get severe. Such measures have likely supported the hesitancy to visit health facilities during the pandemic, which in turn poses a major risk for developing severe malaria (12, 28). The decline observed in March/April 2020 was even more remarkable in inpatients. This does not support our initial hypothesis, that in cases of more severe malaria manifestation, patients were still brought to health facilities and hospitalized, despite the pandemic. The findings from this analysis support the hypothesis, that the reported malaria burden in health facilities will shrink due to the effects of the COVID-19 pandemic in highly malaria-endemic countries (Heuschen et al. 2021). They also support results of the WHO World Malaria Report (12), and they agree with results of similar studies from other SSA countries classified as highly endemic for malaria, such as Sierra Leone, Uganda and the Democratic Republic of the Congo (29-32).

The distinct decrease of OPD visits in the health facilities of northern Ghana in September 2020 may also be explained by unusual heavy floods that started mid-August and could have further complicated the access to health services. Flooded land is a favorable habitat for *Anopheles* mosquitos, the malaria vector, what could have led to the observed increases of malaria incidence in October 2020.

Malaria incidence among pregnant women shows a different trend in northern Ghana. After a decline in reported malaria cases in April 2020, malaria figures have rebounded rapidly in this population and reached even higher levels compared to previous years. The most likely explanation of such an opposite trend would be the hesitancy of pregnant women to visit health facilities. This is probably due to the fear of getting infected with COVID-19, combined with initial disruptions of the provision of intermittent preventive treatment in pregnancy (IPTp) to women in antenatal care (ANC) services as well as the disruption of routine distribution of ITNs (33). The disrupted access to and delivery of ANC services is likely to explain the malaria case trend in April. However, without IPTp and ITNs, more women were at risk for malaria thereafter, which can explain the subsequent rise in malaria cases over the following months. Also, many pregnant women probably have sought the missed ANC after the initial movement restrictions were lifted with subsequent malaria diagnosis.

Ghana had already achieved high levels of ITN coverage, and no ITN mass campaign was planned for 2020 (12). However, the routine distribution of ITNs, which is usually done in health facilities during ANC sessions and in primary schools, needed to be adapted to the COVID-19 measures, which included school closure from March 2020 until January 2021 (34, 35). Also the seasonal malaria chemoprevention intervention for children and the annual indoor residual spraying of insecticides, which both require physical contact between the health workers and the community, needed to be modified (36, 37). As another consequence of the COVID-19 pandemic, the provision of rapid diagnostic tests for malaria is fragile, which may have led to under-diagnosis of cases (38). Finally, reports of hesitancy to visit health facilities due to fear of getting infected with COVID-19 are still common (33, 38). Last but not least, the malaria health care workers capacities were limited due to frequent reassignments to the control of COVID-19, to stigmatization or absence following quarantine, or to the development of COVID-19 disease or even death (13, 35, 39).

This study has strengths and limitations. A strength of the study is that the data represent a whole year of follow-up into the pandemic, which provides a more comprehensive picture of the effects compared to the previous studies with much shorter study periods. Limitations are that the surveillance system itself may have been affected by the pandemic, with a bias in the reported numbers. Moreover, it is not clear if the quality of surveillance data is fully comparable during the five years observed. Finally, much more people with malaria symptoms may have switched to self-medication during the pandemic, which may also have an albeit unknown effect on the malaria figures.

In conclusion, this study shows that the COVID-19 pandemic has been accompanied by a reduced malaria incidence in northern Ghana’s health facilities. Further data from other African countries and in particular data from community-based studies are needed to fully judge the impact of the pandemic on the global malaria situation.

## Data Availability

The datasets used and analyzed in this study are available from the corresponding author on reasonable request.

## Declarations

### Ethics approval and consent to participate

No ethical approval and consent to participate was required as only secondary data have been used.

### Consent for publication

No consent for publication was required (only secondary data used).

### Availability of data and material

The datasets used and/or analyzed in this study are available from the corresponding author on reasonable request.

### Authors’ contributions

AAM and MNA were responsible for the data collection. AH, VW and OR performed the data analysis. AH wrote the first draft under the supervision of OM, AAM and MNA supported the data interpretation. All authors read, reviewed and approved the final manuscript.

## Acknowledgements

We acknowledge financial support by the Else Kröner-Fresenius-Stiftung within the Heidelberg Graduate School of Global Health, by Deutsche Forschungsgemeinschaft within the funding programme Open Access Publishing, by the Baden-Württemberg Ministry of Science, Research and the Arts and by Ruprecht-Karls-Universität Heidelberg.

